# Detecting Self-Repairs from Spontaneous Speech with Prompt Ablation Across LLMs and Fine-Tuned Encoder

**DOI:** 10.64898/2026.08.21.26360471

**Authors:** Rachel Wu, Sydney Pugh, Karen O’Connor, Kevin Xie, Kyra O’Brien, Kevin Johnson

## Abstract

Self-repairs – in-utterance revisions in which a speaker abandons and reformulates their speech – are a promising interpretable marker for speech-based cognitive screening. Detecting them automatically is difficult because a self-repair is defined by its relationship to surrounding speech rather than by fixed lexical cues. On the DementiaBank ADReSS corpus, we compared the capability of generative LLMs under a five-condition prompt ablation against a fine-tuned DistilBERT token classifier at detecting self-repairs. GPT-5 performed best (test F1 = 0.73) and was largely insensitive to prompt design, whereas the LlaMA (open-weight alternative) was both weaker and far more prompt-sensitive (test F1 = 0.47). DistilBERT, nearly 100 times smaller, matched the open-weight LLM at a fraction of the computational cost. These results suggest that a locally deployable encoder, given sufficient in-domain annotation, is a more plausible route to clinical self-repair detection than scaling model size or prompt complexity.

## 1. Introduction

Alzheimer’s disease (AD) is the most common cause of dementia affecting 7.2 million people in the United States over the age of 65.^1^ However, while AD is the most prevalent type of dementia, it accounts for only 60-80% of dementia cases, with the remainder due to other etiologies such as vascular, Lewy body, and frontal-temporal dementias.^1^ These conditions are often grouped together as Alzheimer’s Disease and Related Dementia (ADRD).^2^ Because AD/ADRDs are progressive and currently irreversible, the window in which intervention can meaningfully alter the disease course is early, which makes timely detection critical. Primary care is a natural setting for early detection, as primary care physicians are often the first to encounter patients presenting with cognitive impairment (CI).^3–5^

Early detection of signs of CI allows patients to begin non-pharmacological interventions and disease modifying drug treatments that delay the progression of AD when these interventions are most effective.^6,7^ However, mild cognitive impairment (MCI) is substantially underdiagnosed in primary care with only about 8% of expected cases detected in Medicare patients age 65 and older.^6^ This gap persists because early symptoms are subtle, visit time is limited, and cognitive decline may be difficult to recognize during a routine encounter.^5,6^ These barriers underscore the need for automated tools to help physicians detect signs of CI during the clinical visit.

Subtle alterations in speech fluency have been associated with early changes in cognition.^8–10^ These early indicators, such as prolonged pauses, filled hesitations, and increased disfluency, are often too subtle for standard clinical observation, yet they represent highly quantifiable features for automatic tracking. In our previous study, we presented WATCH-SS: (Warning Assessment and Alerting Tool for Cognitive Health from Spontaneous Speech) a modular, explainable framework that detects signals of CI from a patient’s speech sample.^11^ WATCH-SS combines detectors for five linguistic and acoustic indicators of CI, including filler speech, repetitive speech, substitution errors, vague speech, and speech delays, into a set of clinically interpretable features used for CI classification. On the DementiaBank ADReSS dataset,^12,13^ WATCH-SS achieved strong predictive performance (AUC = 80%) while preserving a transparent diagnostic profile appropriate for primary care screening. Because the framework is modular, individual detectors can be added or refined as additional speech markers of CI are identified.

In the present study, we develop and evaluate methods for detecting an additional marker, self-repairs (revisions), in which a speaker interrupts an utterance in progress to correct or reformulate what they were saying. Self-repairs have been observed to increase in early cognitive decline, where they may reflect greater effort in self-monitoring and word retrieval.^14^ Detecting self-repairs automatically is challenging, however, because a self-repair is defined by its relationship to surrounding speech rather than by fixed lexical cues, and existing approaches have typically relied on detectors trained on annotated conversational corpora, developed to remove disfluencies rather than detect them. Consistent with the design of WATCH-SS, self-repairs are not intended to serve as a standalone marker of CI. Instead, each indicator contributes complementary evidence that is aggregated into an overall risk profile. Our goal in this study is to develop and evaluate methods that accurately detect self-repairs in spontaneous speech, as a step towards incorporating self-repair detection into speech-based cognitive screening tools such as WATCH-SS.

## 2. Related Work

### 2.1. Self-repairs as an indicator of cognitive impairment

Self-repairs and other disfluencies have long been documented in the speech of individuals with AD/ADRD,^10^ where they are thought to reflect underlying production and self-monitoring difficulty.^15,16^ Conversation-analytic and machine learning studies have reported that disfluencies, including fillers and self-repairs, are among the features that help distinguish speech of individuals with AD/ADRD from that of healthy controls. Rohanian et al.^17^ applied a word-level disfluency detector^18^ to tag repair onsets, edit terms and fluent words, then feed these tags into a Bi LSTM model that classifies AD and predicts Mini Mental State Examination (MMSE) scores, showing that these disfluency features improve over word-only models. Using conversational speech from the Carolinas Conversation Collection, Nasreen et al.^19^ found that disfluency features, including self-repairs, contributed to AD classification from spontaneous speech, with the strongest performance achieved by combining disfluencies and interactional features. While much of this evidence comes from individuals already diagnosed with dementia or mild to moderate AD, speech -based markers, including disfluency and related phenomena, have demonstrated diagnostic utility for MCI in a growing body of work, as summarized in a 2025 systematic review and meta-analysis.^14,20^Mueller et al.,^14^ for example, found that in a longitudinal cohort of late middle-aged adults, a connected-language fluency measure that included revisions and false starts declined faster in those with early sub-clinical CI. While no single disfluency feature is diagnostic on its own, within an aggregation-based framework, even modest, complementary signals can strengthen a combined risk profile.

### 2.2. Computational methods to detect self-repairs

Automatic detection of self-repairs has a long history in language processing, grounded in the structural account of disfluencies introduced by Shriberg^21^ who provided a taxonomy and structural model of self-repairs (reparandum-interregnum-repair) that underlies many automatic disfluency and repair detection systems. Across the literature, disfluency detection has traditionally been framed as a preprocessing step with the goal of identifying and removing reparanda to create clean, fluent text for downstream tasks and has used conversational corpora such as Switchboard.^22^ Various methods have been used for repair detection including noisy-channel models,^23^ sequence tagging models that label each token as fluent or disfluent^18^ to BiLSTM models with explicit repair states.^24^ More recently, transformer encoders have been implemented for the task, including self-attention models improved through self-training^25^ and BERT-based detectors adapted for low latency incremental use.^26^

In spite of the progress made using state of the art models, detecting repairs remains difficult for certain repair type. High performance is achieved in the detection of repeated speech (F1 = 0.96) but performance declines in semantically richer repairs most relevant to word finding difficulties, such as substitutions (F1: 0.77-0.79) and deletions and restarts (F1= 0.54), with accuracy further degrading as the repair length increases.^26^

More recently, large language models have been applied to both disfluency detection^27^ and Alzheimer’s detection from spontaneous speech.^28,29^ To date, however, these methods have been developed to remove disfluencies from healthy speech or as an end-to-end classification, rather than to detect and quantify self-repairs as an interpretable marker. In this study, we develop and compare a BERT-based token classification detector and generative large language models (LLaMA and GPT-5), evaluating their ability to detect self-repairs in spontaneous speech from clinically healthy and those with CI as an interpretable feature that can contribute to a clinical risk prediction model.

## 3. Methods

### 3.1. Data

We developed and evaluated methods for detecting self-repairs with the goal of identifying an additional interpretable indicator to contribute to the WATCH-SS framework.^11^ We frame the detection of self-repairs as the identification of repair structures,^21^ and evaluate two modeling approaches: a fine-tuned encoder model and generative large language models (LLMs).

We reuse the ADReSS dataset^12,13^ and pre-processing pipeline from,^11^ consisting of diarized transcripts and audio recordings from participants with and without an Alzheimer’s disease (AD) diagnosis describing the Cookie Theft picture from the Boston Diagnostic Aphasia Examination,^30^ annotated using the CHAT coding system.^31^ We refer readers to^11^ for the full dataset and pre-processing details and wesummarize the key points relevant to the present work below.

The dataset comprises 108 participants for model development and 48 for testing, with all partitions balanced for age, gender, and AD diagnosis. We further split the development-facing partition into training and development (dev) subsets via a stratified 70/30 split, selecting – from 100 candidate splits stratified by gender and diagnosis label – the split minimizing the difference in mean age between the resulting subsets. Table 1 summarizes the resulting partitions. **Data Pre-processing**. As in,^11^ CHAT transcripts were normalized by replacing pause annotations with “[silence]”, event annotations with “[*<* event *>*]” tokens, and inaudible-speech annotations with “[inaudible]”, and removing all other CHAT-specific markup. Only patient-produced utterances were retained for the present analysis.

**Table 1:** Characteristics of the ADReSS train, development (dev), and test datasets.

| Split | Num. Subjects | AD |  | Control |  |
| --- | --- | --- | --- | --- | --- |
| | | Pct. Female | Age (mean ± std) | Pct. Female | Age (mean ± std) |
| train | 75 | 56% | 66.5 ± 6.6 | 55% | 66.6 ± 6.8 |
| dev | 33 | 52% | 67.4 ± 6.7 | 56% | 65.7 ± 6.0 |
| test | 48 | 54% | 66.1 ± 7.4 | 54% | 66.1 ± 7.1 |

The self-repair labeling procedure described below: deriving ground-truth `verdict` directly from CHAT’s `[//]` retracing annotations is introduced in the present work and was not part of the pre-processing pipeline in.^11^

Ground-truth labels were extracted from the original CHAT annotations for all patient utterances. In CHAT, a self-repair is marked with the retracing annotation “[//]” immediately following the reparandum. In cases where the reparandum is a phrase, the full reparandum is delimited by angle brackets (`<>`) in the transcript (e.g., `<the dog is> [//] the cat is running`). CHAT does not separately delimit the boundaries of the corresponding repair. To extend the CHAT annotations, we added labels to all utterances marked with a retracing annotation in the original data set. In these annotations, the repair that follows a marked revision [//] was labeled using a similar convention and the label [rpr]. Single word repairs were labeled with [rpr] (e.g., `a dog [//] beast [rpr]`), and phrase repairs were surrounded with angle brackets (`<>`) and followed by [rpr] (e.g., `<what did you> [//] <how can you>[rpr] see it?`). Annotations were completed by an expert in data annotation with a background in biomedical informatics. All annotations were performed in accordance with the annotation guidelines (Supplemental Material). In total, 264 repair annotations were added to the CHAT reparandum annotations as some utterances had multiple reperandum.

An utterance was labeled positive for self-repair (verdict = `true`) if it contained at least one `[//]` annotation, with the reparandum span taken directly from the text enclosed in `<>`. Extraction of the repair span was instead left entirely to each LLM’s own judgment at inference time, and the verdicts produced by the LLMs were independent of CHAT-derived annotations.

The prevalence of self-repairs across the data splits is summarized in Table 2.

**Table 2:**
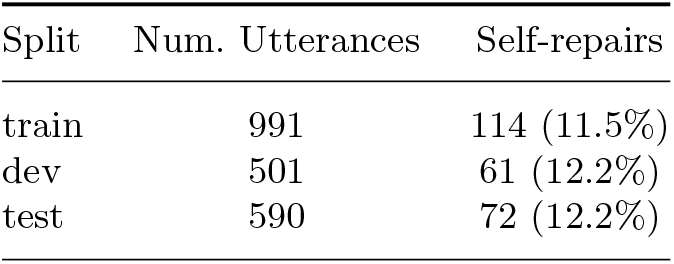
Prevalence of self-repairs in the ADReSS train, development (dev), and test datasets. Values are counts and proportions of positively-labeled utterances, derived from CHAT retracing ([//]) annotations.

| Split | Num. Utterances | Self-repairs |
| --- | --- | --- |
| train | 991 | 114 (11.5%) |
| dev | 501 | 61 (12.2%) |
| test | 590 | 72 (12.2%) |

### 3.2. Prompt Conditions

To assess the contribution of individual prompt-design choices to detector performance, we constructed five prompt conditions as an ablation relative to a *baseline* prompt. Each condition isolates a single design dimension relative to the baseline: the granularity of the exclusion criteria, persona framing, chain-of-thought reasoning, and few-shot demonstrations. All five conditions share the same core operational definition of a self-repair, the criteria and exclusions distinguishing genuine repairs, and the same JSON output schema, differing only along a single dimension as outlined in Table 3. The baseline prompt is summarized in Figure 1, and the complete prompt, along with all prompt conditions, is provided in the supplementary material. The baseline prompt and the varying design dimensions can be summarized as follows:

**Table 3:** Summary of the five prompt conditions used in the ablation. Each condition is a modification of the Baseline prompt along one design dimension.

| Condition | Exclusion detail | Persona framing | CoT reasoning | Few-shot examples |
| --- | --- | --- | --- | --- |
| Baseline | Full (7 items) | No | No | 4 |
| Minimal Exclusions | Reduced (4 items) | No | No | 4 |
| Neurologist Persona | Full (7 items) | Yes | No | 4 |
| Reasoning First | Full (7 items) | No | Yes | 4 |
| Zero-Shot | Full (7 items) | No | No | 0 |

**Fig. 1:**
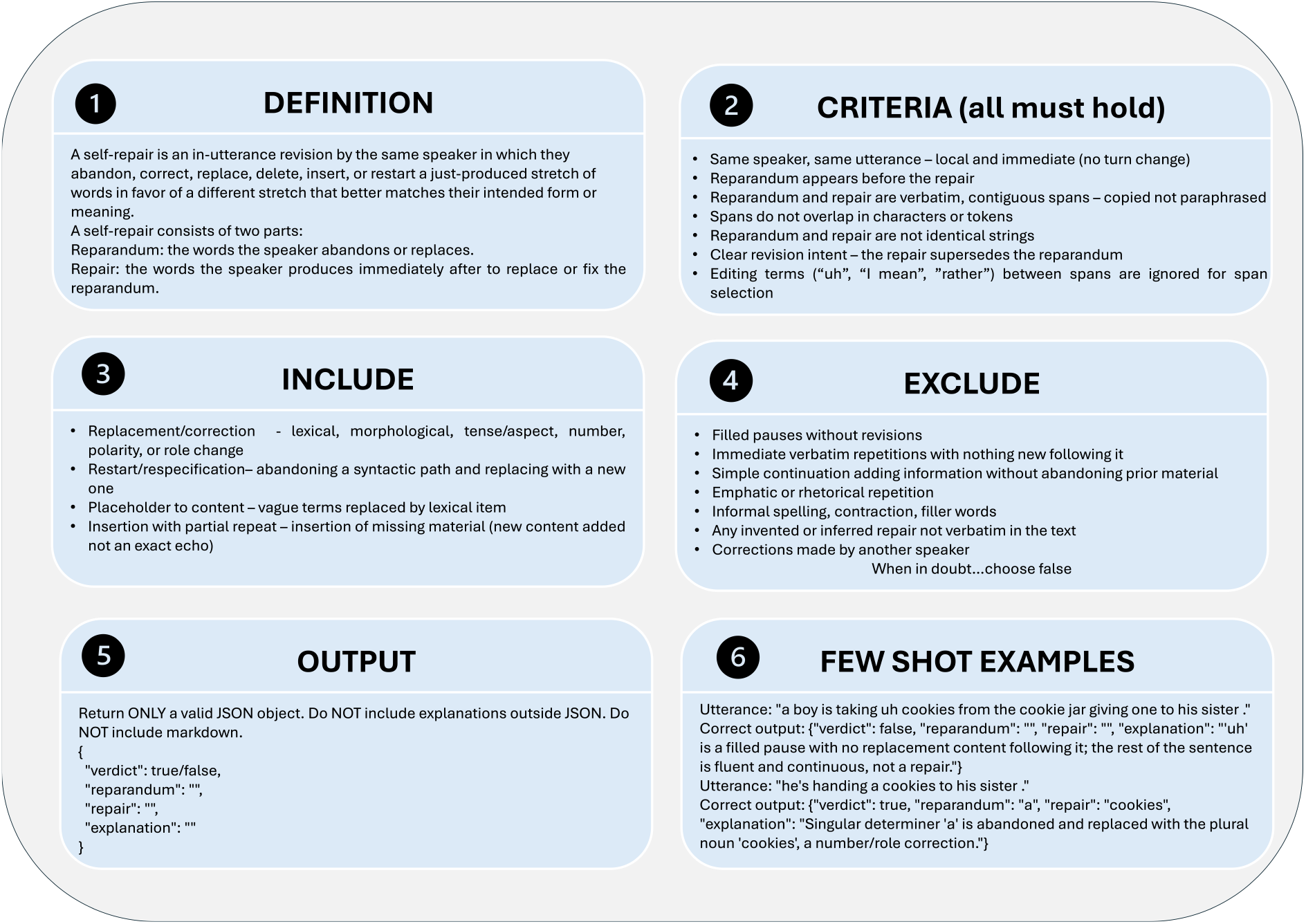
Structure of the baseline self-repair detection prompt, condensed and re-ordered for presentation. The prompt provides a definition of self-repair, criteria that must all hold, categories of repairs to include or exclude, a strict JSON output schema and worked few shot examples (2 of 4 shown). The complete, verbatim prompt and all examples appear in the supplementary material.

- **Baseline**. The full operational definition, a detailed seven-item exclusion list (filled pauses, verbatim-repetition stalls, simple continuation, emphatic repetition, informal register/dialectal variants, invented/uninstantiated repairs, and other-speaker corrections), and four worked examples.
- **Minimal Exclusions**. Identical to Baseline except the exclusion list is reduced to four coarse-grained items, omitting the detailed treatment of verbatim-repetition stalls and informal-register variants as distinct exclusion cases.
- **Neurologist Persona**. Identical to Baseline, prepended with a clinical framing instructing the model to act as a neurologist evaluating a patient’s Cookie Theft picture description for signs of cognitive impairment.
- **Reasoning First**. Identical to Baseline, but the model is instructed to produce a step-by-step `reasoning` field, working through the operational criteria and exclusions, before committing to a `verdict`. The four worked examples are extended with model reasoning traces to demonstrate the expected format.
- **Zero-Shot**. Identical to Baseline (full exclusion list) but with all four worked examples removed, isolating the contribution of few-shot demonstrations.

For every condition, the target utterance was substituted into a fixed *{*`utterance`*}* place-holder at the end of the prompt, and the utterance was scored in isolation, without any preceding or subsequent transcript context.

### 3.3. Generative LLM Models and Inference

We evaluated each of the five prompt conditions against two LLMs spanning a range of scale and openness: GPT-5^32^(OpenAI, accessed via API) and Llama-3.1-8B-Instruct^33^ (Meta, open-weights). Both models were run within the University of Pennsylvania’s HIPAA-compliant Azure environment.

This yields a 5 *×* 3 ablation design (five prompts *×* three models), each applied independently to every utterance in the dataset. To account for generation stochasticity, each (prompt, model, utterance) combination was sampled five times. Temperature was fixed at 1.0 for Llama-3.1-8B-Instruct. GPT-5 does not expose a user-configurable temperature parameter, so default API sampling behavior was used for that model.

For each sample, the model returned a JSON object containing a binary `verdict`, and, when `verdict` was `true`, a `reparandum` span, a `repair` span, and a free-text `explanation` (with an additional `reasoning` field preceding `verdict` for the Reasoning First condition).

### 3.4. Post-hoc Hallucination Filtering

Because LLMs can produce reparandum/repair spans that are plausible but not actually grounded in the source utterance, a post-hoc hallucination filter was applied to every positive (`verdict = true`) model output before scoring. For each candidate (reparandum, repair) pair, the filter checked two criteria against the corresponding utterance:

1. **Verbatim-in-text:** both the reparandum and repair spans must occur, under loose normalization (lowercased, whitespace-collapsed, punctuation-stripped), as substrings of the utterance.
2. **Non-identical:** the reparandum and repair must differ under normalization (case- and whitespace-insensitive); identical spans do not constitute a genuine repair.

A candidate was retained only if it passed both checks (strict conjunctive filtering); failing either check caused the corresponding utterance’s detector score to be revised from 1 to 0, regardless of the model’s original verdict.

### 3.5. False-Positive Rate as a Function of Utterance Length

Beyond aggregate precision and recall, we examined whether false positives are systematically associated with utterance length. Longer utterances offer more opportunities for verbatim or near-verbatim word recurrence, hedges, and syntactic restarts that superficially resemble self-repair without meeting our operational criteria, so we hypothesized that false-positive rate (FPR) would increase with utterance length.

#### Length-bucketed FPR

For each (model, prompt) condition, we restricted analysis to ground-truth negative utterances (no annotated self-repair) and grouped them into six word-count buckets: 1–5, 6–10, 11–15, 16–20, 21–25, and 26+. Because each condition was sampled five times, we pooled predictions across reruns at the utterance-rerun level: for a bucket containing *N* negative utterances, the denominator was 5*N* negative utterance-rerun instances, and the numerator was the total count of those instances for which the model’s (post-filter) verdict was a false positive. Bucket-level FPR is thus FPR_*b*_ = FP_*b*_*/*(5*N*_*b*_).

### 3.6. Encoder-Based Baseline: DistilBERT Token Classification

In addition to evaluating how well LLMs could identify self-repair and spans, we were also interested in examining if smaller encoder language models could perform comparably. Said encoder models could have significant utility as low-cost, low-compute solutions for real-time processing of text. Given the small size of our annotated dataset, we chose DistilBERT,^34^ a 67-million parameter model as our test case. We framed self-repair identification as a token classification task and finetuned DistilBERT for either 3-label (Outside, Repair, Reparandum), and 5-label (Outside, Begin-Repair, Inside-Repair, Begin-Reparandum, Outside-Reparandum) classification. In line with Soni and Roberts^35^, we first finetuned DistilBERT using the Switch-board dataset, which was similarly labeled for self-repair^36^, with a 98-2 train-validation split. We then further finetuned DistilBERT on our annotated dataset with a 80-20 train-validation split. To measure the variability of the encoder model, we repeated this experiment 10 times, with different randomly generated train-validation splits. We further conducted an ablation study, removing the intermediate finetuning on the Switchboard dataset, to determine its impact on model performance. All encoder experiments were performed in Python with the Huggingface Library.

## 4. Results

### 4.1. Prompt Ablation Performance

We ran all LLM experiments five times for each prompt condition. The result reported for each prompt is the mean of those five runs. Tables 4 and 5 report the performance of the five prompt conditions for self-repair detection on the ADReSS training data, for GPT-5 and Llama-3.1-8B-Instruct, respectively. The results show a clear distinction based on model scale and openness. GPT-5 achieves strong, stable performance across all five prompt conditions, while the open-weights model shows much lower absolute performance and far greater sensitivity to prompt design.

**Table 4:** GPT-5 Prompting Strategy Comparison for Speech Self-Repair Token Classification (5 runs per condition)

| Prompt Type | F1 | Precision | Recall | Accuracy |
| --- | --- | --- | --- | --- |
| **Baseline** | **0.7775 ± 0.0117** | **0.6784 ± 0.0103** | 0.9105 ± 0.0144 | **0.9400 ± 0.0032** |
| Minimal Exclusions | 0.7707 ± 0.0195 | 0.6608 ± 0.0228 | **0.9246 ± 0.0118** | 0.9366 ± 0.0062 |
| Neurologist Persona | 0.7707 ± 0.0120 | 0.6618 ± 0.0131 | 0.9228 ± 0.0200 | 0.9368 ± 0.0035 |
| Reasoning First | 0.7722 ± 0.0286 | 0.6735 ± 0.0322 | 0.9053 ± 0.0273 | 0.9385 ± 0.0085 |
| Zero Shot | 0.7651 ± 0.0104 | 0.6703 ± 0.0132 | 0.8912 ± 0.0100 | 0.9370 ± 0.0033 |

**Table 5:** Llama-3.1-8B-Instruct Prompting Strategy Comparison for Speech Self-Repair Token Classification (5 runs per condition)

| Prompt Type | F1 | Precision | Recall | Accuracy |
| --- | --- | --- | --- | --- |
| Baseline | 0.3748 ± 0.0081 | 0.3038 ± 0.0093 | 0.4895 ± 0.0114 | 0.8121 ± 0.0058 |
| **Minimal Exclusions** | **0.3867 ± 0.0101** | 0.3252 ± 0.0111 | 0.4772 ± 0.0118 | 0.8258 ± 0.0055 |
| Neurologist Persona | 0.3864 ± 0.0186 | **0.3924 ± 0.0176** | 0.3807 ± 0.0211 | 0.8610 ± 0.0040 |
| Reasoning First | 0.3336 ± 0.0144 | 0.2174 ± 0.0104 | **0.7175 ± 0.0227** | 0.6700 ± 0.0120 |
| Zero Shot | 0.3018 ± 0.0270 | 0.4462 ± 0.0334 | 0.2281 ± 0.0223 | **0.8787 ± 0.0037** |

**GPT-5** achieved the the highest performance across all prompt conditions with mean *F*_1_ varing by only 1.2 points across the entire ablation (76.5%–77.8%). The Baseline prompt performed best (mean *F*_1_ = 77.8%, precision = 67.8%, recall = 91.1%). Recall remains consistently high (89–92%) regardless of prompt condition, while precision is the binding constraint on *F*_1_ throughout, indicating GPT-5’s errors are dominated by over-prediction rather than missed detections.

**Llama-3.1-8B-Instruct** achieves its best mean *F*_1_ under Minimal Exclusions (0.3867 *±* 0.0101) and its lowest under Zero-Shot (0.3018 *±* 0.0270), a much wider 0.0849-point spread than GPT-5’s. The best performer is Minimal Exclusions, achieving a modest mean *F*_1_ of 38.7%, shortly followed by Neurologist Persona (38.6%). These two conditions, however, trade off differently, with Neurologist Persona raising precision to 39.2% while achieving the lowest recall among the top three conditions (38.1%). Reasoning First and Zero-Shot mark the two failure extremes. Reasoning First achieves the highest recall (0.7175) but the lowest precision (0.2174) and the lowest accuracy overall (0.6700), indicating the chain-of-thought instruction increases over-triggering rather than improving judgment. Zero-Shot achieves the highest precision (0.4462) but the lowest recall (0.2281) and lowest mean *F*_1_ (0.3018), suggesting that the model defaults to conservative predictions without worked examples to anchor its judgment.

### 4.2. False-Positive Rate Increases with Utterance Length

Table 6 reports length-bucketed FPR (Section 3.5) for GPT-5 (baseline prompt) against Llama (`minimal_exclusions`, window off vs. on); Table 7 reports the fitted slope and *R*^2^ for all prompt conditions. GPT-5’s FPR grows only weakly and non-monotonically with length (*R*^2^ = 0.68 for the baseline prompt, dipping at the 16–20 word bucket), whereas Llama-3.1-8B-Instruct shows a substantially stronger and more consistent trend across all prompt conditions.

**Table 6:** False-positive rate (FPR) by utterance-length bucket, pooled across five inference reruns. GPT-5 (baseline prompt) shows no consistent length trend; Llama (best config, minimal_exclusions) shows a clear increase.

| Length (words) | GPT-5 (baseline) | Llama |
| --- | --- | --- |
| 1–5 | 1.1% | 2.0% |
| 6–10 | 4.7% | 12.8% |
| 11–15 | 16.4% | 27.2% |
| 16–20 | 7.9% | 30.5% |
| 21–25 | 12.0% | 45.3% |
| 26+ | 20.0% | 54.3% |

**Table 7:** Log-linear fit of log(FPR) on utterance length (bucket midpoint, in words) for every prompt configuration, plus mean F1 (5 reruns) for reference. A larger slope/*R*^2^ indicates a stronger, more consistent exponential relationship between length and false positives.

| Model | Prompt config | Slope | $R^2$ | Mean F1 |
| --- | --- | --- | --- | --- |
| GPT-5 | baseline | 0.093 | 0.68 | 0.777 |
| GPT-5 | `minimal_exclusions` | 0.099 | 0.72 | 0.771 |
| GPT-5 | `neurologist_persona` | 0.088 | 0.65 | 0.771 |
| GPT-5 | `reasoning_first` | 0.090 | 0.65 | 0.772 |
| GPT-5 | `zero_shot` | 0.118 | 0.72 | 0.765 |
| Llama | baseline | 0.110 | 0.61 | 0.375 |
| Llama | `minimal_exclusions` | 0.113 | 0.78 | 0.387 |
| Llama | `neurologist_persona` | 0.217 | 0.69 | 0.386 |
| Llama | `reasoning_first*` | 0.045 | 0.33 | 0.334 |
| Llama | `zero_shot` | 0.188 | 0.94 | 0.302 |

\*Non-monotonic (FPR falls at the 26+ bucket), giving a weak fit; excluded from the length-trend claim in the text.

Llama’s `reasoning_first` condition is non-monotonic (FPR falls at the 26+ bucket) and is excluded from the length-trend claim, though its raw fit is reported in Table 7 for completeness.

### 4.3. Test-Set Evaluation

Table 8 reports performance on the held-out ADReSS test set for each model’s best training-set configuration: GPT-5 with Baseline, and Llama-3.1-8B-Instruct with Minimal Exclusions. The two models diverge in how their test performance relates to training: GPT-5’s test F1 is modestly lower than its training F1 (77.8% *→* 72.7%), consistent with a small degree of overfitting to the prompt tuned on the training split, while precision, recall, and accuracy all declined (precision: 67.8% *→* 62.9%; recall: 91.1% *→* 86.1%; accuracy: 94.0% *→* 92.1%). Despite this, GPT-5’s test performance remains well above Llama’s in absolute terms across every metric.

**Table 8:** Test-Set Performance: GPT-5 and Llama-3.1-8B-Instruct (5 runs per condition)

| Model | Prompt Type | F1 | Precision | Recall | Accuracy |
| --- | --- | --- | --- | --- | --- |
| GPT-5 | Baseline | $0.7269 \pm 0.0132$ | $0.6291 \pm 0.0162$ | $0.8611 \pm 0.0098$ | $0.9210 \pm 0.0046$ |
| Llama-3.1-8B-Instruct | Minimal Exclusions | $0.4683 \pm 0.0062$ | $0.3901 \pm 0.0037$ | $0.5861 \pm 0.0181$ | $0.8376 \pm 0.0022$ |

By contrast, Llama’s test performance, interestingly, is modestly higher than its training performance on every metric (F1: 38.7% *→* 46.8%; precision: 32.5% *→* 39.0%; recall: 47.7% *→* 58.6%; accuracy: 82.6% *→* 83.8%).

### 4.4. Encoder-Based Baseline Performance

Table 9 summarizes DistilBERT’s token classification performance across label schemes and pretraining conditions, averaged over 10 runs. Intermediate finetuning on Switchboard substantially improved performance, raising mean *F*_1_ from 0.2557 to 0.4042 for the 3-label scheme and from 0.2126 to 0.3792 for the 5-label BIO scheme. The simpler 3-label scheme modestly outperformed the 5-label BIO scheme under both conditions.

**Table 9:** Experiment Comparison: Speech Self-Repair Token Classification (DistilBERT-base-uncased, 10 runs per condition)

| Label Scheme | Switchboard | F1 | Precision | Recall | Accuracy |
| --- | --- | --- | --- | --- | --- |
| 3-label | Yes | $0.4042 \pm 0.0456$ | $0.4108 \pm 0.0527$ | $0.3999 \pm 0.0489$ | $0.8123 \pm 0.0193$ |
| 5-label BIO | Yes | $0.3792 \pm 0.0470$ | $0.3726 \pm 0.0518$ | $0.3890 \pm 0.0558$ | $0.7951 \pm 0.0194$ |
| 3-label | No | $0.2557 \pm 0.0375$ | $0.2559 \pm 0.0410$ | $0.2568 \pm 0.0406$ | $0.7829 \pm 0.0166$ |
| 5-label BIO | No | $0.2126 \pm 0.0378$ | $0.2247 \pm 0.0414$ | $0.2032 \pm 0.0392$ | $0.7649 \pm 0.0223$ |

## 5. Discussion

Across the LLM models evaluated, GPT-5 achieved the strongest utterance-level self-repair detection, *F*_1_ of approximately 0.73 on the test set, compared with 0.47 for Llama-3.1-8B-Instruct. Although GPT-5 was the clear performance leader, several factors limit its use in real clinical screening settings. Transmitting patient speech to a closed, externally hosted model raises privacy and audibility concerns, API-based inference at scale would carry recurring per-use cost, and although the model returns the reparandum and repair spans, it’s decisions are not interpretable.

These constraints point toward a self-contained application that runs it models locally, or within a controlled environment, rather than calling an external service. The current performance ceiling and cost structure associated with GPT-5 are, however, unlikely to be permanent. Increasingly capable models are being released rapidly, inference costs have fallen consistently, and open-weight models have repeatedly approached the capability of leading closed models within short windows. It is therefore reasonable to expect that a model with GPT-5 level performance will become cheaper, locally deployable, or available as open weights, will be available. In the meantime, GPT-5 provides a strong reference point for the performance achievable for this task.

A central finding is that model size did not straightforwardly determine performance. Despite being nearly 100 times smaller, the fine-tuned DistilBERT performed comparably to Llama3.1-8B. DistilBERT’s performance increased by roughly 0.15 *F*_1_ when Switchboard data was included, suggesting that its performance may currently be more limited by the scarcity of in-domain labeled data than by model capacity. The open-weight LLMs showed a similar pattern. Across five prompting configurations of increasing sophistication, limited improvement in performance was produced, indicating that the gap to GPT-5 was not primarily a prompt design problem but a capability limitation at this model scale. Closing the gap may require parameter-efficient fine tuning (e.g., Low-Rank Adaptation (LoRA)) on annotated repair data rather than further prompt engineering.^37^

This interpretation is consistent with prior work in clinical NLP, where domain adapted and fine-tuned models have increased performance on specialized tasks,^38,39^ and where fine-tuned smaller encoder models have matched or exceeded large language models. Xie et al. found that a prompt-engineered Llama2-13B model fell short of a finetuned BioClinicalBERT model when classifying if patients were having seizures or were seizure free in epilepsy clinical notes. Ojemann2025-vi Similarly, Chang et al. found that a finetuned BERT model was comparable to both zero- and few-shot DeepSeek-8B model in three-way classification of patient epilepsy and seizure types.^40^ While comparable in performance, smaller encoder models require significantly less resources and can run one or even two orders of magnitude faster.^40,41^ This suggests that a fully finetuned encoder model could be deployed in edge-computing environments that would preclude effective use of a large language model, making them an attractive alternative for clinical screening applications. Additional labeled data, either from gold-standard manual annotations or distilled from premiere large language models, could push DistilBERT’s performance even higher.

One bottleneck to performance on smaller models was the size of the available annotated data set. LLaMA 3 8B was the natural open-weight, locally deployable candidate, but performance was roughly the same as BERT and well below GPT-5.

### 5.1. Limitations and Future Work

This study relied solely on the ADReSS dataset and its existing annotations, and no independent inter-annotator agreement was established for self-repairs. Self-repairs are defined relationally and can be genuinely ambiguous to annotate, so the reference labels themselves carry uncertainty that places an unknown ceiling on achievable *F*_1_. Our reported scores should be read against that ceiling rather than against perfect ground truth. Evaluation was limited to picture-description speech from a single corpus, which constrains generalizability beyond this task, and evaluation is only at the utterance level, which does not directly measure span-level accuracy.

DistilBERT operated at the utterance level, and self-repairs are defined by their relationship to surrounding speech. Extending the input to include neighboring utterances may give the model cross-utterance information a token-level model currently lacks. We plan to incorporate this in future work. The underlying data limitation could also be addressed by enlarging the annotated repair data for training. One route for this is synthetic augmentation, which has improved disfluency detection in prior work,^42,43^ though care must be taken to ensure that the generated repairs reflect the characteristics of those produced in cognitive decline rather than generic conversational disfluencies. A complementary approach is a shared, commonly annotated benchmark. Much of the progress in speech-based cognitive assessment have come through shared tests such as ADReSS, which provided balanced, commonly annotated data against which methods can be compared directly. A shared benchmark specifically annotated for self-repairs in clinical speech does not currently exist. Building such a corpus, would not only establish inter-rater reliability, but would likely accelerate progress more than model changes alone.

## 6. Conclusion

We developed and evaluated methods for detecting self-repairs in spontaneous speech as a candidate additional indicator for the WATCH-SS framework. A large generative model (GPT-5) detected self-repairs most accurately, but its cost, opacity, and reliance on external hosting limit its suitability for clinical deployment, while a compact fine-tuned encoder (DistilBERT) matched a far larger open-weight model at a fraction of the computational cost. Across models, performance was limited less by scale than by the scarcity of annotated repair data, pointing to in-domain fine-tuning and shared annotated resources as a promising path forward. These results support self-repair as a viable additional signal for an interpretable, aggregate risk profile, and identify a clear and achievable set of next steps toward integrating it into WATCH-SS.

## Data Availability

The ADReSS dataset is available from the DementiaBank database at https://talkbank.org/dementia/ADReSS-2020/index.html. The reannotated Switchboard Corpus can be found at https://github.com/vickyzayats/switchboard_corrected_reannotated.

https://github.com/vickyzayats/switchboard_corrected_reannotated

## Funding Statement

This work was supported by the National Institutes of Health, United States [DP1LM014558]

## Notes

### Competing Interest Statement

The authors have declared no competing interest.

### Author Declarations

The study used (or will use) ONLY openly available human data that were originally located at the DementiaBank database at https://talkbank.org/dementia/ADReSS-2020/index.html and the Switchboard corpus at https://github.com/vickyzayats/switchboard_corrected_reannotated

